# CuNA: Cumulant-based Network Analysis of genotype-phenotype associations in Parkinson’s Disease

**DOI:** 10.1101/2021.08.02.21261457

**Authors:** Aritra Bose, Daniel E. Platt, Niina Haiminen, Laxmi Parida

## Abstract

Parkinson’s Disease (PD) is a progressive neurodegenerative movement disorder characterized by loss of striatal dopaminergic neurons. Progression of PD is usually captured by a host of clinical features represented in different rating scales. PD diagnosis is associated with a broad spectrum of non-motor symptoms such as depression, sleep disorder as well as motor symptoms such as movement impairment, etc. The variability within the clinical phenotype of PD makes detection of the genes associated with early onset PD a difficult task. To address this issue, we developed CuNA, a cumulant-based network analysis algorithm that creates a network from higher-order relationships between eQTLs and phenotypes as captured by cumulants. We also designed a multi-omics simulator, CuNAsim to test CuNA’s qualitative accuracy. CuNA accurately detects communities of clinical phenotypes and finds genes associated with them. When applied on PD data, we find previously unreported genes INPP5J, SAMD1 and OR4K13 associated with symptoms of PD affecting the kidney, muscles and olfaction. CuNA provides a framework to integrate and analyze RNA-seq, genotype and clinical phenotype data from complex diseases for more targeted diagnostic and therapeutic solutions in personalized medicine. CuNA and CuNAsim binaries are available upon request.

## 1 Introduction

A primary goal in complex disease genetics is to understand how genes influence the symptoms, that is, the mapping from genotype to phenotype. The knowledge about etiology and pathogenesis of a disease provides a basis for targeted treatment and prevention. Casecontrol genome wide association studies (GWAS) and whole-exome sequencing (WES) are useful methods to understand the rare causative mutations that underlie complex diseases with small effects from common variants [1]. Quantitative Trait Loci (eQTL) studies bridge these methods by enabling investigation of the effect of the genotypes or risk loci on gene expression levels and how, in turn, they affect phenotypes [2]. expression eQTL analysis is used to determine hotspots, construct causal networks, discover stratification in clinical phenotypes and select genes for clinical trials [3]. The application of these methods have revealed a significant number of risk loci [4–6] in complex diseases.

Parkinson’s Disease (PD) is such a complex neurological disorder affecting approximately 1.2% of the world’s septuagenarian population. PD has a rapid progression characterized by motor symptoms due to loss of dopaminergic neurons in the substantia nigra and presence of Lewy bodies [7], bradykinesia, rigidity and tremor [8]. PD progresses from early symptoms such as mild non-motor manifestations to significant degenerative effects on mobility and muscle control [9] in advanced stages. The progression of symptoms of PD is tracked by rating scales which asses different stages of the disease. The most widely accepted rating scale is the Hoehn and Yahr (HY) scale [10], while another comprehensive assessment scale is the Unified Parkinson’s Disease Rating Scale sponsored by the Movement Disorder Society (MDS-UPDRS) [11]. Recently, over 41 genetic susceptibility loci have been associated with late-onset PD in the largest GWAS meta-analysis up to date [12]. Few genes have been found to be causal among these risk loci, but for majority of loci, it is not yet known which genes are linked with PD risk. Moreover, despite concerted efforts in understanding the genomic processes underlying the progression of the disease, the clinical heterogeneity of PD makes it elusive. There is a complex interaction between motor and non-motor symptoms, with both impacting key issues such as sleep, constipation, depression and muscle movement [13, 14]. It has also been hypothesized that PD actually comprises two subtypes, brain-first or body-first [15]. Due to this heterogeneity in clinical features and their trajectories, it is important to understand the biological processes underlying these groups of features and symptoms.

To this end, we developed CuNA, namely, Cumulant-based Network Analysis. CuNA finds higher order genotype-phenotype interactions by integrating genes implicated in the disease as obtained from GWAS or eQTL studies and the associated phenotypes or clinical features. Hence, we find groups of features from the similar subsets of subjects using logical relationships among features called “redescription” clusters and subsequent cumulant computations. CuNA performs community detection on the network constructed from the significant higher-order interactions between clinical features and genes related to the disease. To show that CuNA accurately captures the interaction between the biomarkers and phenotypes, we designed CuNAsim, a simulator for gene expression, genotypes and phenotypes. CuNAsim is a multi-omics simulator which simulates genomics and transcriptomics data accounting for endophenotypes. It also captures eQTLs and relationships between omics data with an array of clinical phenotypes. Although in framework it is similar to a prior multi-omics simulator [16], CuNAsim provides simulation scenarios with relative correlation of each phenotype with a user defined set of biomarkers (genes and genotypes).

To disentangle the effects of heterogeneity of PD, we applied CuNA to the collection of data from the Parkinson’s Progression Markers Initiative (PPMI) study (https://www.ppmi-info.org). We found several novel genes associated with a collection of PD phenotypes. Although previous work has demonstrated success in predicting PD status from gene expression data (e.g. [17]), associations of genes with the phenotypic measurements underlying PD diagnosis have not been reported before at this level of granularity. CuNA enables us to find such interactions which are often not captured by traditional GWAS, highlighting the clinical heterogeneity of the disease. Although, we apply CuNA to understand the biological underpinnings of motor and non-motor symptoms of PD, the method can be applied to a host of complex diseases which are captured by an array of clinical features, symptoms, environmental and behavioral effects such as Alzheimer’s Disease, Coronary Artery Disease ad metabolic syndrome, Cancer and other neurological disorders. CuNA finds biomarkers associated with these clinical and non-genetic features paving the path for future biomarker discovery and therapeutics for complex diseases.

## 2 Methods

### 2.1 CuNAsim

CuNAsim is a multi-omics simulator integrating phenotypes, genotypes, and gene expression levels. To handle the integration of different omics data we started with a multivariate distribution

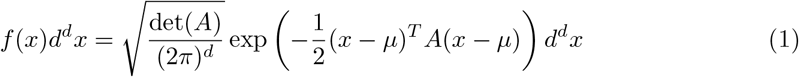

Components of *x* were identified as phenotypic (binary, which may include environmental conditions as well), SNP (pairs of binary alleles, one for each of the chromosome pairs), or gene expression (floating). Covariances *A*^−1^ were specified in terms of *A* = *σ*cor(*x, x*^*T*^)*σ* where the *σ* is a diagonal matrix with values representing the spread of the variates, and cor(*x, x*^*T*^) is specified to yield correlations among phenotypes, alleles between each pair of chromosomes representing Hardy-Weinberg disequlibrium, and among gene expression levels reflecting coregulation among pathways. Correlations between phenotypes, SNPs and expression levels reflect interactions including allele impacts on expression levels, relationships between SNPs, expression levels, and disease/phenotype processes, driven by biological pathways. Offsets *µ* set quantities such as MAF, case/control proportions, and expression level centers. Binary values were mapped from *I*(*x*_*i*_ ≥ 0). The fraction of cases are *E*(*I*(*x*_*i*_ ≥ 0)). Genotypes were mapped from *I*(*x*_*I*_ ≥ 0) + *I*(*x*_*i*+1_ ≥ 0). MAF is then *E*(*I*(*x*_*i*_ ≥ 0)). ORs may be derived from the joint probabilities *E*(*I*(*x*_*i*_ ≥ 0 ∧ *x*_*j*_ ≥ 0)) for SNP values *x*_*i*_. Expression levels were mapped to exp(*x*_*i*_).

We simulated three different simulation scenarios for 1,000 samples and 11 features (3 phenotypes, 3 SNPs and 5 genes with varying expression levels). Although CuNAsim can generate high dimensional data we restricted our toy simulation to demonstrate the accuracy of CuNA in picking out the genotype-phenotype interactions with the highest Pearson correlation coefficient (*r*^2^) and to demonstrate its robustness in presence of false positives and correcting for spurious associations. To achieve this objective we designed three scenarios with varying correlations. In the first scenario, we designed an extreme case where only a few features among the genes, SNPs and phenotypes were highly correlated with each other (inset in Figure 2). In the second case, we took an average case where many of the features were moderately correlated with each other (Supplementary Figure 5). For the third case we performed a sanity check with completely uncorrelated features, therefore, the resulting correlation matrix being equal to an identity matrix.

### 2.2 Parkinson’s Disease Data

#### RNA-seq expression data

We compiled RNA-seq gene expression data from the PPMI phase 2 release containing 4,649 blood-based samples across five visits and 34,386 genes with Transcripts per million (TPM) values. PPMI annotates samples with labels reflecting whether they are from de novo PD subjects (subjects diagnosed with PD for two years or less and are not taking PD medications; annotated as PD) and from control subjects without PD who are 30 years or older and do not have a blood relative with PD diagnosis (annotated as HC). We used PD (n=293) and HC (n=163) samples only from the baseline visit for our analyses as the number of overlapping samples with genotype and gene expression data for other visits were low.

#### Genotype data

The genotype data released in Phase 1 of PPMI contained 960 individuals and approximately 44 million high quality Single Nucleotide Polymorphisms (SNPs) that passed GATK VQSR quality control. We further filtered SNPs with missing genotyping rate *>* 0.02 for SNPs and individuals, respectively and Minor Allele Frequency (MAF) *>* 0.05, Hardy-Weinberg equilibrium *>* 1*e* − 6 and removed individuals with heterozygosity rates with more than three standard deviations from the mean resulting in 5.6 million SNPs. We only selected PD and HC samples having baseline gene expression data, 456 individuals.

### 2.3 CuNA

CuNA integrates the phenotypes related to PD (or any disease) along with the genetic variants or genes as features, and computes higher-order associations between these features to find subsets of features influencing groups of individuals with similar underlying biological pathways. An outline of the algorithm is given in Algorithm 1. CuNA computes cumulants and construct networks with only statistically significant connections between any two pair of features *i* and *j*. It computes *N*_*i,j*_ as a tuple of number of cumulant groups containing both *i* and *j* denoted as *n*_*i,j*_, number of cumulant groups containing only *i*: *n*_*i*,*_, number of cumulant groups containing only *j*: *n*_*,*j*_ and number of cumulant groups without either of *i* or *j*. This allows us to compute a Fisher’s exact test and obtain significance parameters for each pair *i* and *j* and whether the edge between them in a network would be at random. We form the network with pairs of features which has a *p <* 0.05 in the Fisher’s exact test.

#### Algorithm 1 CuNA: Cumulant-based Network Analysis

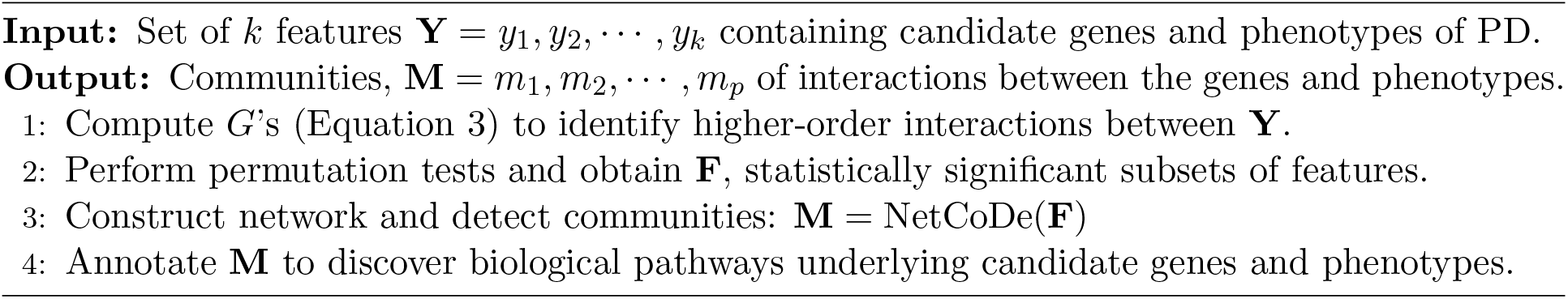

#### Algorithm 2 NetCoDe: Network formation and community detection

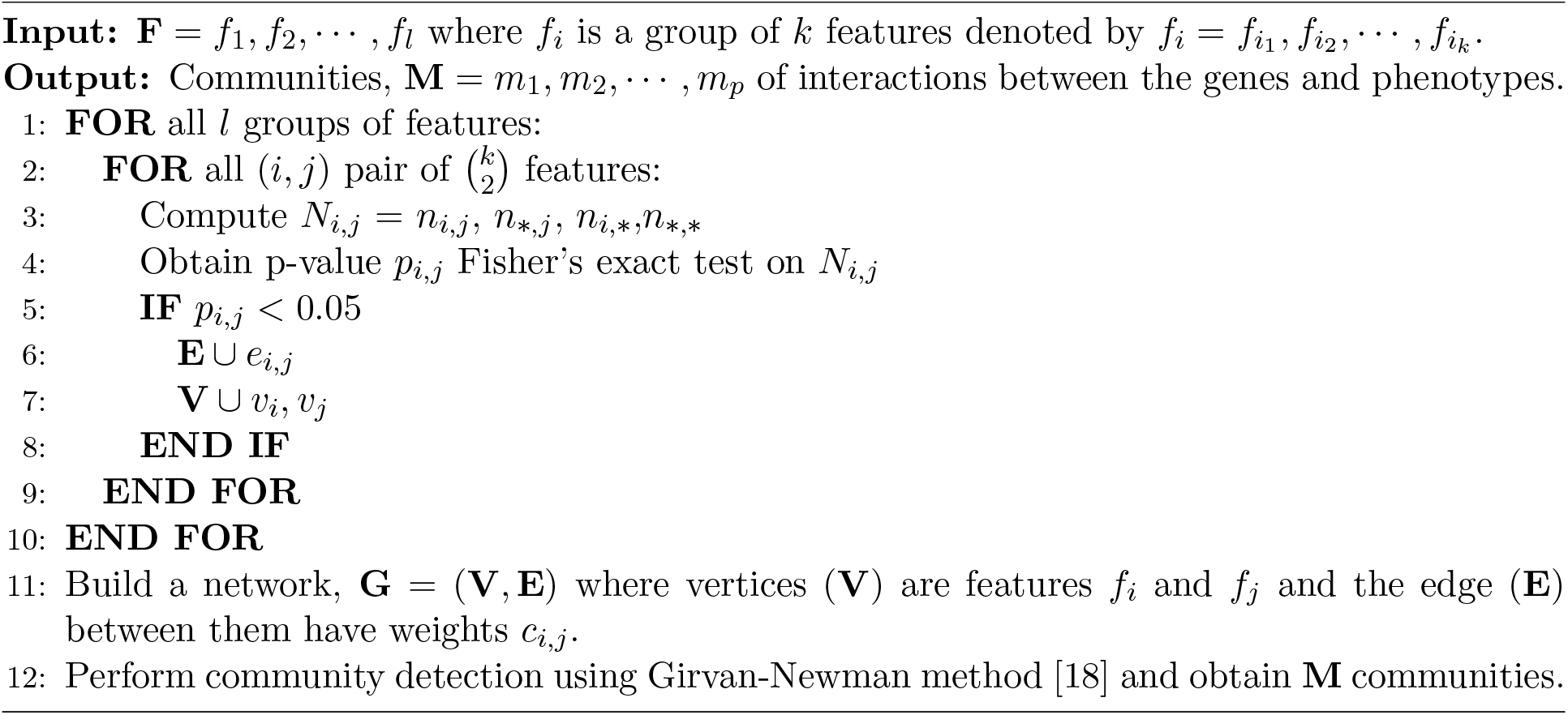

### 2.4 Study design

CuNA is a framework to study the genetic factors influencing the clinical features of a complex disease, in this case, PD with its motor and non-motor symptoms. As a first step, we take the genotype data as well as the RNA-seq gene expression data as input and compute eQTLs. We extract significant *cis-eGenes* (above a predefined statistical significance threshold) and include them as features with the phenotypic measurements related to PD. Thereafter, we apply CuNA (Cumulant-based Network Analysis) as a meta-analysis method on these candidate genes and phenotypic features in order to draw higher-order associations between them. We construct a network as part of CuNA and perform community detection on the network to obtain communities or clusters of interacting features (genes and phenotypes). Further gene ontology analysis is performed on these interacting genes to obtain the biological pathways highlighted for similar symptoms or features in PD. The outline of our approach is detailed in Figure 1.

**Figure 1:**
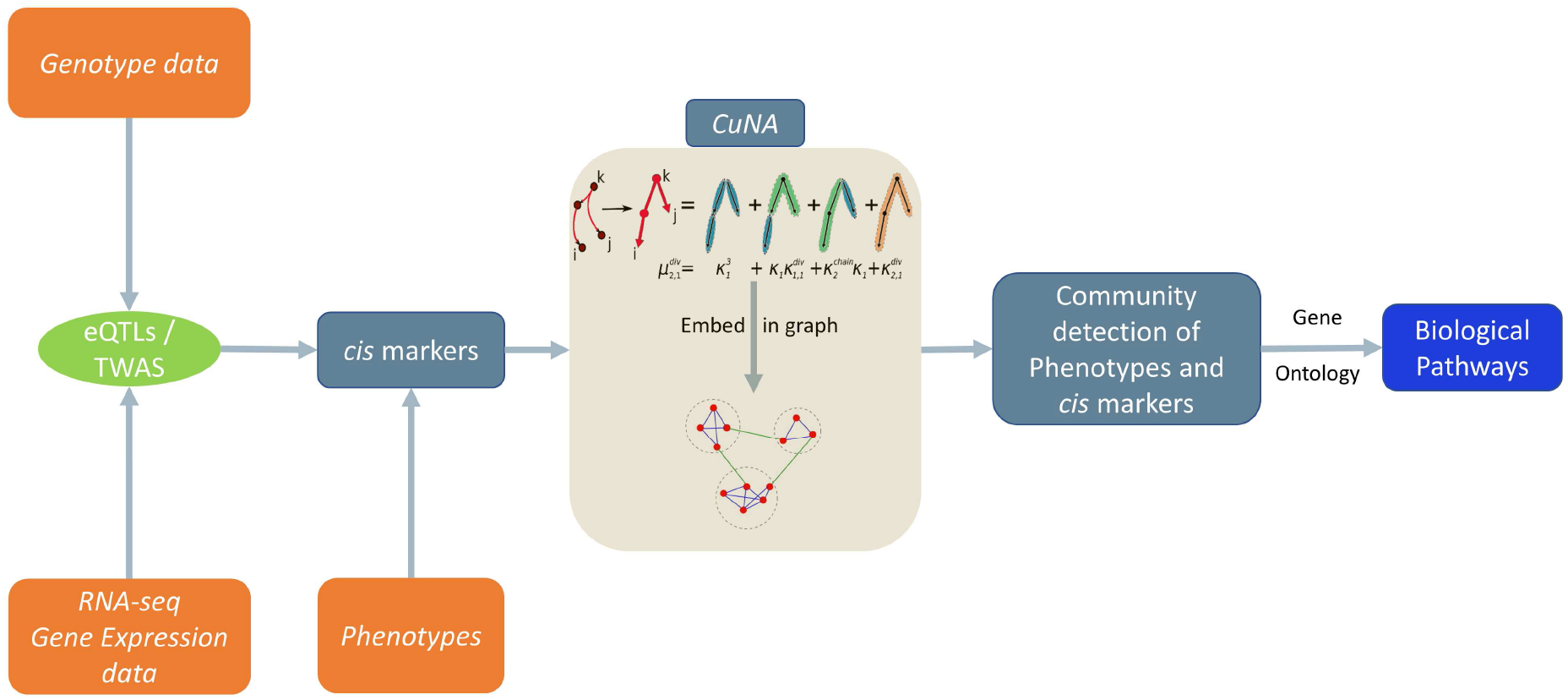
Overview of the study design with CuNA playing a central role. Inputs are colored in orange and output in blue.

#### Computing eQTLs

We used Matrix eQTL [19] for fast eQTL analysis on 34,386 genes and 5.6 million SNPs across 456 individuals. For all of our eQTL analysis we used *p*-value threshold of 1 × 10^−7^ and FDR *<* 0.05 and a distance of 1 × 10^6^ base pairs in which the gene-SNP pair would be considered local and tagged as *cis*-eQTL (Supplementary Figure 1). Matrix eQTL tests for association between each SNP and transcript by modeling the effect of genotype as either additive linear or categorical. We computed the top 20 Principal Components (PCs) of the genotype data using TeraPCA [20] and included them along with age and gender information as covariates to correct for latent population structure (Supplementary Figure 2).

#### Supervised classification

We used machine learning approaches from Python’s scikit-learn 0.23.2 package to classify HC from PD on 456 individuals (293 PD and 163 HC), with 25% of the data used for validation. We applied the Synthetic Minority Oversampling Technique (SMOTE) [21] to balance the PD and HC classes as we have more cases than controls. We used a host of classifiers such as Random Forest, Linear Regression, Ridge Regression, Support Vector Machine (SVM) with linear and Radial Basis Function (RBF) kernels, etc. on the training data set and performed fivefold cross validation (CV) for finding optimal hyper-parameters. We performed permutation tests using scikit-learn’s model selection for classification to obtain statistical significance (*p*-value) of the performance of the chosen classifier using CV. Once these subsets of features are identified, we obtain statistical significance of each such group by permutation tests and from the significant subsets of features (*p <* 1*e* − 6, FDR*<* 0.05 and |*Z*| *>* 3), we construct a network.

CuNA builds the networks between the features and the edge weights between any two feature representing the number of times these features have grouped together in all the subsets of features in the cumulant computation. The interaction network thus can be very dense with a total of 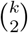 edges with *k* features. We thus allow only a small percentage of edges until we have observed all *k* features due to ease of visualization and analysis. On this network, we perform community detection using the algorithm described in Algorithm 2 and analyze each such community drawing latent interactions between genes and the symptoms or clinical features of the disease.

### 2.5 Cumulants

We seek to identify distinct groups of individuals whose pattern memberships may give hints to underlying pathways involved with disease processes. Relationships between the roles of these features defining the patterns are revealed in how multiple patterns capture the same groups of subjects, called redescriptions (Details in Appendix A). Since most of the progression markers collected in the PPMI are strongly correlated, and we need to factor out those strong lower-order correlations from higher order associations marking distinct groups of individuals differentiating disease processes as their Parkinson’s advances.

One approach towards such a factorization is suggested through a convergence of a number of fields of study. Correlation expansions emerge naturally in quantum field theory, expressed as a series of Feynman diagrams. These factored moments, essentially higher-dimensional cumulants, may be factored to represent a set of “one-particle-irreducible” (1PI) diagrams [22]. Such emerge naturally in statistics of large deviations through Craméssr’s theorem [23], which also connects to the notion of “effective actions” from quantum field theory. Their generating functions satisfy useful set partition relationships, and have been a part of traditional statistics for some time [24].

This factorization is represented by a moment generating function

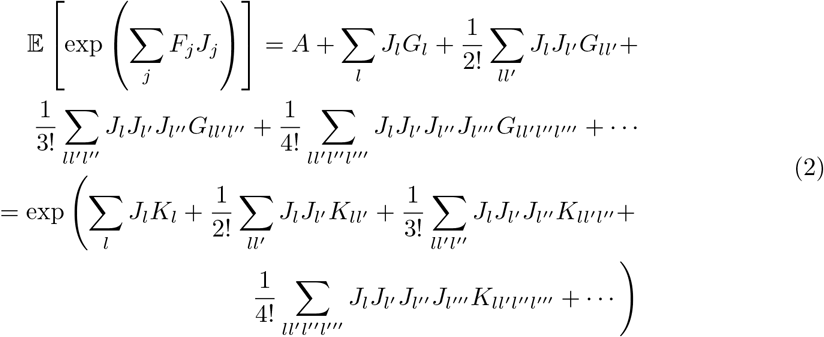

where the *F*_*j*_ are features indexed by *j* (defined in Algorithm 1), the *G*’s represent moments, *A* is a constant offset (unity in this case) defined by *J* = 0, and the *K*’s represent higher order cumulants, e.g. *G*_*ij*_ = *E*(*x*_*i*_*x*_*j*_) and 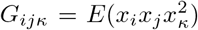, and the *K*_*ij*_ and *K*_*ijκ*_ would be the corresponding cummulants. These may be extracted in terms of the power series to yield

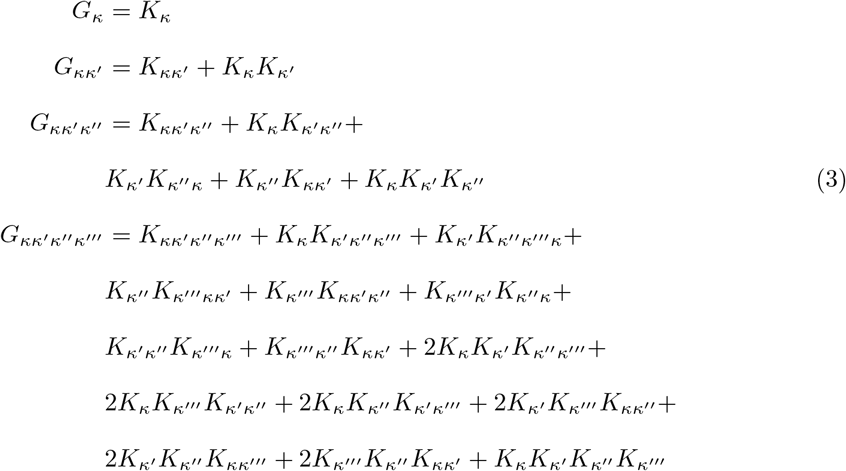

We apply this factorization to patterns, and test significance constructing null hypotheses and variances by shuffling phenotypes.

### 2.6 Redescription clusters

Subjects *s* ∈ 𝒮 are described by a list of features *f*_*i*_(*s*) indexed by feature labels *i* ∈ ℱ. Each feature has an alphabet 𝒜_*i*_ so that *f*_*i*_(*s*) ∈ 𝒜_*i*_ which is often binary, but could be defined on the reals. Examples of binary features in ℱ are diagnoses (Dx) such as PD or other motor and non-motor, symptoms, blood pressure, etc. which would have a continuum alphabet (𝒜_*bmi*_ = ℝ).

For a given *a*_*i*_ ∈ 𝒜_*i*_, the set of subjects that have that value is 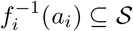. So the list of subjects with PD can be written 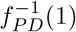. In the case of continuous variables, the selection of sets is according to a threshold, such as the mean *m*(*f*_*i*_(*S*)), mapped to 1 if *f*_*i*_(*s*) ≥ *m*(*f*_*i*_(*S*)).

Patterns may be described in terms of conjunctions *i*∧*j* for *i, j* ∈ ℱ such that 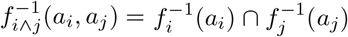 for binary *a*_*i*_, *a*_*j*_. This definition is extended to include either atomic *i, j*, such as PD or T2D, or to any combinations of conjunctions subject to the logical algebra of ∧ (e.g. (*i* ∧ *j*) ∧ (*i* ∧ *k*) = *i* ∧ *j* ∧ *k* for *i, j, k* ∈ ℱ subject to values *a*_*i*_, *a*_*j*_, *a*_*k*_. So we can specify the PD subjects with a motor or non-motor symptom such as walking or handwriting as 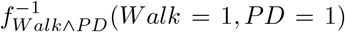. Such combinations of conjunctions *i* that have more or less members 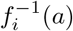 than expected by chance are called patterns.

Binomial and other tests of the significance of patterns can be dominated by lower-order correlations among the variables in a pattern. Two distinct patterns that yield the same subsets of subjects, e.g. 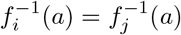, are called “redescriptions.” If conjunctions yield a form such as *A* ∩ *B* = *B*, then it may be deduced that *B* ⊂ *A*, and the conditions yielding *A* and *B* satisfy *b* ⇒ *a*. In other words, redescriptions can reveal logical relationships among features. Such relationships may reflect underlying biological pathways reflected in these connected phenotype patterns. Therefore, each of these patterns *i* specify a phenotype, which may be associated with genotypes or other -omic data using standard methods.

Given the presence of misclassifications, differential evolution of disease stages, simple transcription mistakes, etc, result in errors in estimates of 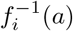 must be accounted for in estimating equivalence. We can use Jaccard distances 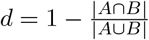 measures deviations. So 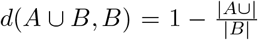 is 0 if *B* ⊆ *A*, some non-zero value with any *B* ⊄ *A*. This distance measures the probability that samples drawn from *A* and *B* are not shared, which gives an index for the possible to distinguish disruption due to errors, or whether it would be possible to distinguish non biological pathways vs. biological pathways with error.

### 2.7 Pathway Analysis

We performed gene ontology by doing pathway enrichment analysis of the 24 cis-genes using the package clusterProfiler 3.8 [25] in Rwith the KEGG database [26], with *p <* 0.05 and Benjamini-Hochberg false discovery rate adjustment.

## 3 Results

### 3.1 Simulation study

We applied CuNA on the data simulated by the multi-omics simulator CuNAsim which was developed particularly for integrating genomics, transcriptomics and phenotypes. In the first scenario we allowed only a few highly correlated interactions between the features such as (*Gene0* — *SNP0*) with *r*^2^ = 0.9, (*Gene0* — *SNP2*) with *r*^2^ = 0.8, (*Pheno0* — *Pheno1*) with *r*^2^ = 0.6, etc. Using the simulated data from the first scenario as an input to the CuNA pipeline, we found the resulting embedded network from higher order interactions captures all the aforementioned interactions (Figure 2). Running the community detection algorithm on the network (Figure 2) we found the following communities:

**Figure 2:**
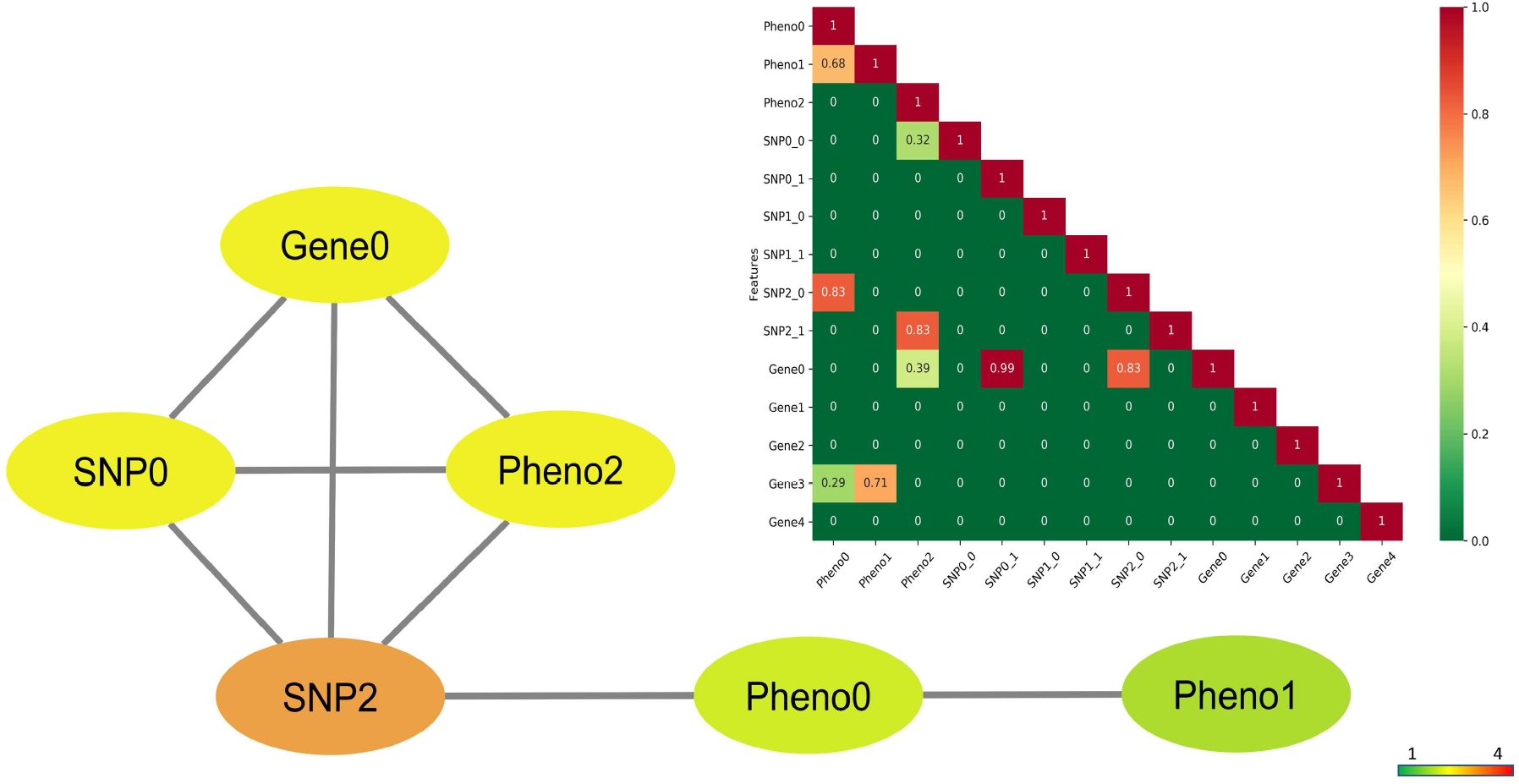
Network of the simulated variables from the first scenario with highly correlated features. The nodes are colored by degrees (darker colors have higher degree). The correlation matrix of the variables is shown in the inset with the color gradient.

- *Gene0, SNP2, Pheno2*
- *SNP0*
- *Pheno0, Pheno1*

Thus, CuNA captures the communites accurately as reflected in the network (Figure 2) as well as the original correlations which was the input to Algorithm 1.

For the average case with moderately correlated interactions between features as shown in Supplementary Figure 5, we see a similar behavior when we applied CuNA. It captures the highly correlated interactions such as (*Gene0* — *SNP1*), (*Gene0* — *Gene1*), (*Gene2* — *Pheno1*), etc. The communities also reflected clusters of biomarkers and phenotypes which followed the input correlation matrix as shown in the inset of Supplementary Figure 5. They were:

- *Gene0, Gene1, Gene2, SNP1, Pheno2*
- *SNP0, Pheno0*
- *Pheno1*

For another extreme case of no correlation between the features we found none of the interactions crossed our user defined threshold of *p <* 1*e* − 6, |*Z*| *>* 3 and FDR*<* 0.05. As expected, CuNA failed to observe anything significant from uncorrelated features even when we increased the parameters for checking false positive associations. The parameters for generating the simulated correlations with mean *µ* and standard deviation *σ* for each feature is detailed in Supplementary Tables B-D for the first simulation scenario and in Supplementary Tables E-G for the second scenario.

### 3.2 Selecting predictive cis-eGenes

We computed eQTLs on the 456 PD and HC individuals having genotype and gene expression data from the baseline visit. We obtained 24 *cis* and 53,550 *trans* significant SNP-gene pairs. Given that *trans*-eQTL analyses are more prone to be affected by systematic errors between genomic regions than *cis*-eQTLs [27], we only considered *cis-eGenes*. Several of the associated *cis-eGenes* play a functional role in PD and are found to be significant in GTEx v8 analyses, expressed in brain tissues [28]. The *cis-eGenes* include known PD-associated genes such as the ubiquitin ligase NEDD4 which is protective against *α*−synuclein accumulation and toxicity in animal models of PD [29], AGO2 which co-participates with PD gene LRRK2 [30], KIF1A which is a key regulator of neural circuit deterioration in aging leading to intellectual disability, muscle weakness, etc. [31], and LRTM1 whose cells survive and differentiate into midbrain dopaminergic neurons *in vivo* resulting in significant improvement in motor behavior [32]. In addition, several of the genes are known to be expressed in the brain but not previously implicated in PD. Details about the protein-coding *cis-eGenes* and their expression in brain and other tissues can be found in Supplementary Table A.

We evaluated the performance of the 24 *cis-eGenes* in disease classification with machine learning methods on the blood-based gene expression data. The best performing method on the 75% training set was SVM with RBF kernel (Supplementary Figure 4). SVM (RBF kernel) resulted in an *F*_1_ score of 0.61 with precision and recall of 0.62 and 0.65, respectively on the 25% test set. This result was statistically significant (permutation test *p*-value 0.009). When we applied the SVM classification using all the genes in the RNA-seq data, we observed a similar *F*_1_ score of 0.62 as well as similar precision (0.63) and recall (0.66) on the test set (Table 1). Hence, the 24 *cis-eGenes* preserve the performance of the entire set of 34,386 genes when classifying PD cases vs. healthy subjects.

**Table 1:** Classification performance of the 24 *cis-eGenes*, compared to using all genes.

| Status | Precision | Recall | $F_1$ score |
| --- | --- | --- | --- |
| *cis-eGenes* HC | 0.53 | 0.24 | 0.33 |
| *cis-eGenes* PD | 0.67 | 0.88 | 0.76 |
| *cis-eGenes* Total | 0.62 | 0.65 | 0.61 |
| All genes Total | 0.63 | 0.66 | 0.62 |

KEGG pathway enrichment analysis of the 24 *cis-eGenes* revealed one statistically significant pathway, inositol phophate metabolism (Supplementary Figure 6). Phosphatidylinositol 4,5 biphosphate enhances the presence of *α*-synuclein’s membrane association [33]. Inositolphosphate signaling pathway may act to reduce autophagy and in turn play a vital role in neurodegeneratie diseases [34] such as PD (due to a decline in autophagy).

### 3.3 CuNA reveals genotype-phenotype relationships

We combined for CuNA the gene expression data on the *cis-eGenes* and the motor and non-motor phenotypes obtained from the PPMI study which included the MDS-UPDRS features, HY scale, age, sex, etc. We computed the cumulants to find higher-order interactions between all the features (including *cis-eGenes*). The cumulants’ ability to separate higher-order moment contributions from possibly strong lower-order terms is highly desirable, and shows separability when applied to PPMI data contrasted with binomial tests of pattern significance.

Starting from 15,275 sets of features with similar patterns we filtered for significance by applying a threshold for *p <* 1*e* − 6 and FDR*<* 0.05 and obtained 761 significant sets of features. We constructed the network of dense interactions among all the associated features from these sets. The gene SAMD1 and the MDS-UPDRS variable *NP2SWAL* (chewing and swallowing issues) play central roles in the network with the top 20% of the edges (Supplementary Figure 3). Allowing more edges make the network denser and does not add new nodes (features). Hence, for visualization purposes we use the top 20% of the edges.

To disentangle the interactions between genes and PD phenotypes we performed community detection on the network (Supplementary Figure 3) and obtained the following community clusters:

- A cluster with variables *Dx* (diagnosis) and *NHY* (Hoehn-Yahr scale).
- A second cluster with the variable *NP2SWAL* playing a central role with other non-motor symptoms such as *NP2SALV* (saliva and drooling) and *NP2SPCH* (speech). Other features such as *NP1FATG* (fatigue), *NP1LTHD* (light headedness) and *NP1SLPD* (day-time sleepiness) also interact in this cluster.
- A third cluster with the gene SAMD1 interacting with MDS-UPDRS variables such as *NP1PAIN* (pain), *NP1WALK* (walking and balance), *NP1CNST* (constipation) and *NP1URIN* (urinary problems). Genes such as INPP5J and OR4K13 along with the phenotype *Olfact* are also present.

For visualizing the communities in detail, we computed the Maximum Spanning Tree (MST) (Figure 3) of the entire network (Supplementary Figure 3).

**Figure 3:**
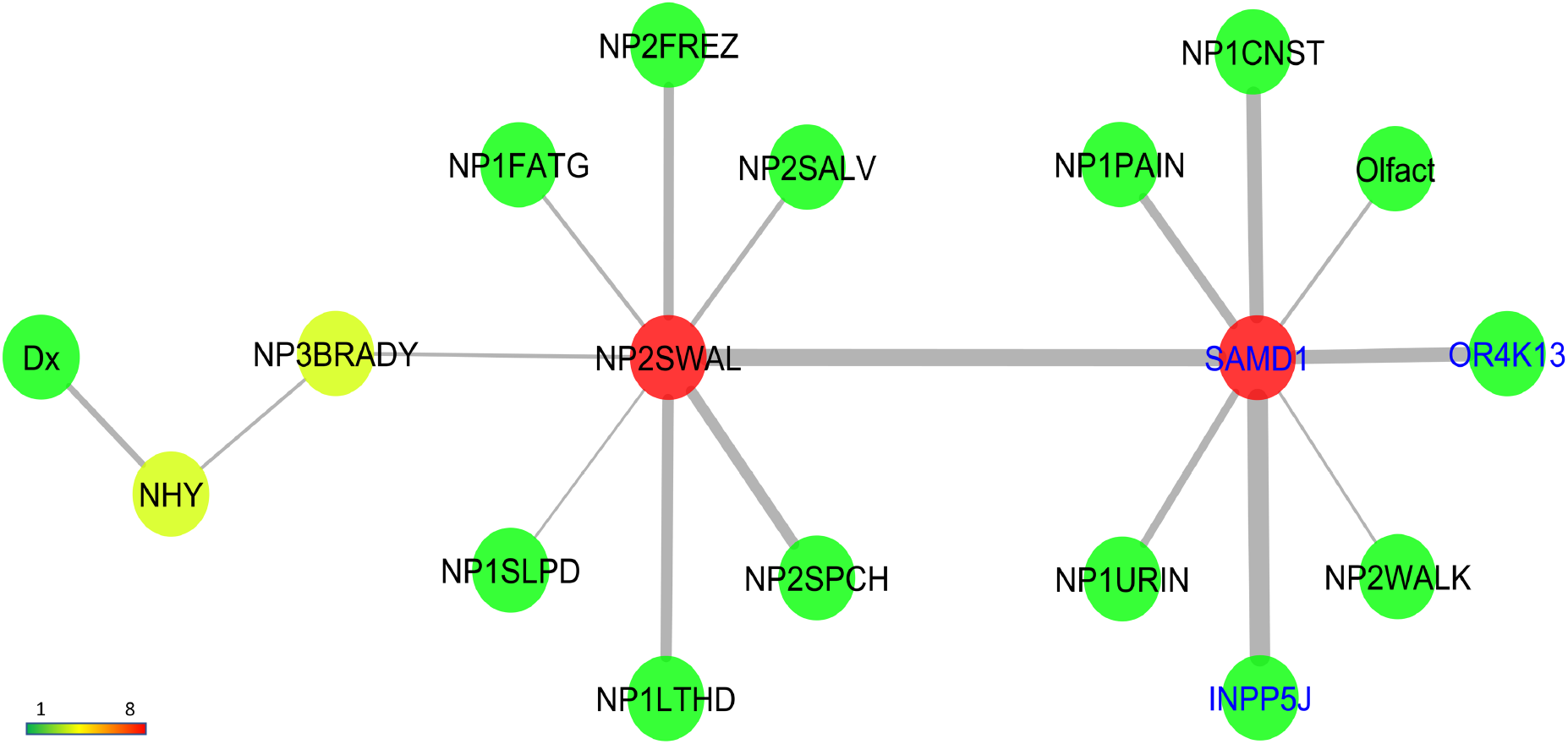
Maximum Spanning Tree (MST) of the network generated by embedding higher-order cumulants. Nodes representing phenotypes and genes have black and blue labels, respectively. Edge widths are directly proportional to their weights. Color of nodes relate to their degree, red being the highest and green being lowest.

The first community with the variables *Dx* and *NHY* are straightforward to interpret as PD diagnosis and HY scale are instrumental tools to understand the disease progression. The second community contains MDS-UPDRS variables which are all related to the movement of mouth muscles with saliva, drooling, speech and swallowing. Also present are variables related to dizziness and fatigue such as light-headedness, daytime sleepiness, etc. which are all early symptoms of PD. Lastly, and most importantly, we observe the gene SAMD1 plays a very crucial and central role in the network. SAMD1 is expressed in blood and immune system including T lymphocytes as well as in brain tissues. T lymphocytes have been shown to recognize *α*−synuclein peptides in PD patient [35] and thus we present evidence for a previously unreported association of the gene SAMD1 in PD diagnosis and early symptoms. The gene INPP5J is also present in the cluster and is known to be associated in Lowe syndrome which causes renal failure and affects the brain. Here, likewise, it interacts with MDS-UPDRS variables related to constipation and urinary problems in early onset PD. Also present in the cluster is the gene OR4K13 (Olfactory receptor gene) which interacts with the phenotype *Olfact* capturing olfactory problems in early onset PD patients. Thus, CuNA reveals the relationships with genes and clinical features of PD as represented by MDS-UPDRS variables, HY scale, etc. decoding the heterogeneity of the clinical features of PD.

## 4 Discussion

The cumulant-based network analysis, CuNA, introduced here, can be used to detect genes associated with clinical features in higher-dimensional space, adding granular view in contrast to traditional case-control studies. There is a dearth of methods addressing the genetic associations and underlying biological pathways of the symptoms and clinical features of idiopathic PD or other complex diseases. This approach provides a framework integrating genotype, gene expression and endophenotypes as input and finds relationships between them. eQTLs and genotype-phenotype interactions are often plagued by false positives due to uncorrected confounding effects such as population structure, environmental factors, etc. Hence, it is important to test the robustness of CuNA to find whether it captures true biomarkers associated with phenotypes of interest. We designed CuNAsim, a fast and efficient multi-omics simulator which supports an array of phenotypes or clinical features to be tested alongside genotypes and gene expression data. The “piped” algorithm structure of CuNA takes in input the simulated data from CuNAsim and accurately captures the correlated features in forms of communities in the network. Thus, CuNA is robust under different simulation scenarios and accurately finds true associations.

CuNA computes cumulants in the form of redescription groups. Cumulants are higher-order moments and thus expensive to compute. Higher-order cumulants play an important role in the analysis of non-normally distributed multivariate data and the computational complexity increases with the order by a factor of *n*^*d*^, where *d* is the order of the cumulant and *n* is the number of marginal variables. In genomics parlance, this creates a computational bottleneck as the number of variables are in the order of hundreds of thousands with the decreasing cost of sequencing. Thus CuNA undergoes a computational bottleneck in the cumulant computation with increasing number of variables. Advances in randomized algorithms and tensor decomposition allows for faster computation of cumulants. A possible future direction is to make CuNA faster by leveraging super-symmetric tensors in block structures and efficient cumulant computation.

Applying CuNA to a Parkinson’s disease data set of genotype and RNA-seq expression data from blood samples with associated multitude of phenotypic measurements, we found several novel candidate genes associated with PD phenotypes. We run CuNA on the candidate significant *cis-eGenes* obtained by computing eQTLs. These *cis-eGenes* captured similar case-control classification performance as the whole data set. They were also enriched in the inositol phosphate metabolism pathway which is linked with neurodegenerative diseases such as PD. Thus, the *cis-eGenes* have both biological and statistical significance in the context of PD. As latent population stratification can lead to spurious eQTLs and confound the study, we included the top twenty PCs as covariates in the analysis. CuNA reveals cliques associated with related biological functions such as constipation, urination and renal failure and the gene INPP5J which is implicated in Lowe Syndrome and is found to be significant in both brain and kidney cortex tissues in GTEx analysis (Supplementary Table A). MDS-UPDRS measures were found to be associated with genes such as SAMD1 which is expressed in blood and immune system as well as brain tissues. Blood-based gene expression such as analyzed here has shown similarity with brain-based expression and is an intriguing noninvasive option for capturing neurodegenerative disease progression [36].

CuNA can disentangle the complex higher-order genotype-phenotype interactions, embed them in a network and analyze it. Network analysis and community detection approaches provide a deeper understanding of association studies involving eQTLs and phenotypes of interest with a visualization tool. The hyper-parameters and user-defined parameter thresholds can be varied to observe robustness and sensitivity of the method in handling false positives.

## 5 Conclusion

Associations between genotype, gene expression and phenotype data can be complex and often confounded by various environmental factors. We propose a novel framework CuNA to identify associations with more granularity than a standard case-control association study. We demonstrate that CuNA captures true associations by applying it on simulated data as obtained from our novel multi-omics simulator CuNAsim. When applied to PD diagnostic data encompassing clinical features along with motor and non-motor symptoms, CuNA identifies novel gene-phenotype relationships while replicating already known associations with PD.

GWAS has the potential to find loci with common genetic variants contributing to disease risk. It has been extensively used in PD finding genes associated with disease risk. However, in progressive diseases such as PD, Alzheimer’s, cancer, cardiovascular diseases, etc. with a rich repository of phenotypes or clinical features, it is of significance to study the genes associated with an ensemble of the features sharing similar biological pathway. CuNA provides an exciting opportunity to decode phenotypic and genotypic diversity and discover genes associated with various manifestations of complex diseases, paving the way for future biomarker discovery and personalized therapeutics.

## Data Availability

Data is available through Michael J. Fox Foundation upon request.

## 6 Acknowledgements

Data used in the preparation of this article were obtained from the Parkinson’s Progression Markers Initiative (PPMI) database (www.ppmi-info.org/data). For up-to-date information on the study, visit www.ppmi-info.org.

## Supplementary Materials

**Figure 4:**
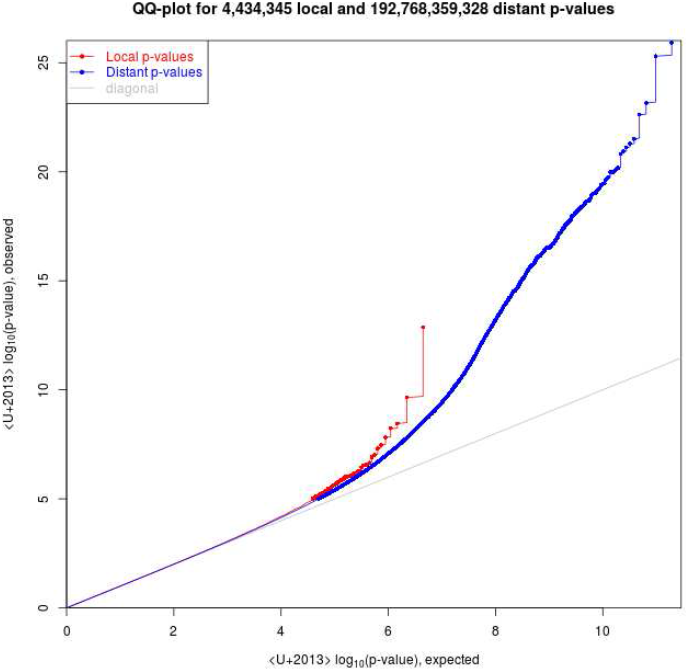
QQ plot showing statistical significance of *cis* (Local p-values, red) and *trans* (Distant p-values, blue) eQTLs.

### eQTL analysis

The goal of eQTL analysis is to identify SNPs which are significantly associated with expression of known genes. They reveal complex biological processes underlying diseased systems and help discover latent genetic factors causing certain diseases. Most eQTL studies perform separate association tests for each transcript-SNP pair. Association testing can be done in a straightforward manner by linear regression ar ANOVA models and if required, non-linear techniques such as generalized linear and mixed models, Bayesian regression [37], accounting for pedigree [38], etc. Many methods have been developed to find groups of SNPs associated with expression of a single gene [8, 39]. With the advancement of sequencing techniques and decreasing cost there has been an unprecedented growth in genotype and expression level data. As eQTL studies identify SNPs which are significantly associated with expression of known genes, they can be computationally intensive resulting in billions of associations for large scale data. The simple linear regression is one of the most commonly used methods for eQTLs.

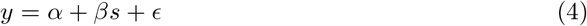

where ϵ ∼ i.i.d *N* (0, *σ*^2^). The number of such tests can easily result in billions. Instead, if we let **G** is the gene expression matrix, with each row containing measurements for a single gene across individuals and **S** be the genotype matrix, with each row containing measurements for a single SNP across individuals. Then the matrix of all gene-SNP correlations can be calculated in one large matrix multiplication. Thus we have,

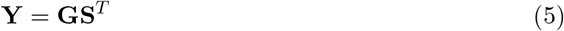

The correlations are thus computed in Equation 5 and we report the corresponding test statistic, *p*-value, FDR, etc.

### Redescriptions

Subjects *s* ∈ 𝒮 are described by a list of features *f*_*i*_(*s*) indexed by feature labels *i* ∈ ℱ. Each feature has an alphabet 𝒜_*i*_ so that *f*_*i*_(*s*) ∈ 𝒜_*i*_. That alphabet is often binary, but could be defined on the reals. Examples of binary features in ℱ are diagnoses (Dx) such as hypertension (HT) or type-II diabetes (T2D), or body mass index (bmi) which would have a continuum alphabet (𝒜_*bmi*_ = ℝ).

For a given *a*_*i*_ ∈ 𝒜_*i*_, the set of subjects that have that value is 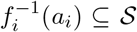. So the list of subjects with hypertension can be written 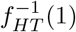. In the case of continuous variables, the selection of sets is according to a threshold, such as the mean *m*(*f*_*i*_(*S*)), mapped to 1 if *f*_*i*_(*s*) ≥ *m*(*f*_*i*_(*S*)).

Patterns may be described in terms of conjunctions *i*∧*j* for *i, j* ∈ ℱ such 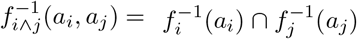 for binary *a*_*i*_, *a*_*j*_. This definition is extended to include either atomic *i, j*, such as HT or T2D, or coronary artery disease (CAD), or to any combinations of conjunctions subject to the logical algebra of ∧ (e.g. (*i* ∧ *j*) ∧ (*i* ∧ *k*) = *i* ∧ *j* ∧ *k* for *i, j, k* ∈ ℱ subject to values *a*_*i*_, *a*_*j*_, *a*_*k*_. So we can specify the diabetic hypertensive subjects as 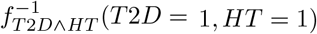. Such combinations of conjunctions *i* that have more or less members 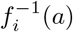 than expected by chance are called patterns.

Binomial and other tests of the significance of patterns can be dominated by lower-order correlations among the variables in a pattern.

Two distinct patterns that yield the same subsets of subjects, e.g. 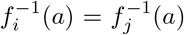, are called “redescriptions.” If conjunctions yield a form such as *A* ∩ *B* = *B*, then it may be deduced that *B* ⊂ *A*, and the conditions yielding *A* and *B* satisfy *b* ⇒ *a*. In other words, redescriptions can reveal logical relationships among features. Such relationships may reflect underlying biological pathways reflected in these connected phenotype patterns. Therefore, each of these patterns *i* specify a phenotype, which may be associated with genotypes or other -omic data using standard methods.

Given the presence of misclassifications, differential evolution of disease stages, simple transcription mistakes, etc, result in errors in estimates of 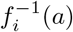 must be accounted for in estimating equivalence. We can use Jaccard distances 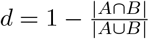 measures deviations. So 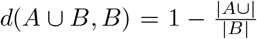 is 0 if *B* ⊆ *A*, some non-zero value with any *B* ⊄ *A*. This distance measures the probability that samples drawn from *A* and *B* are not shared, which gives an index for the possible to distinguish disruption due to errors, or whether it would be possible to distinguish non biological pathways vs. biological pathways with error.

### Population Structure

The PPMI data set has population structure which may confound the eQTL computation and therefore result in spurious associations in downstream CuNA computations. We observe a main cluster of “RAWHITE” which relates to the Europeans and Caucasian ethnicities present in the data set. Another cluster appears in the scatterplot of the top two PCs (Figure 5) related to the “RABLACK” or the African ethnicities in the data. These legends are defined by the PPMI study.

**Figure 5:**
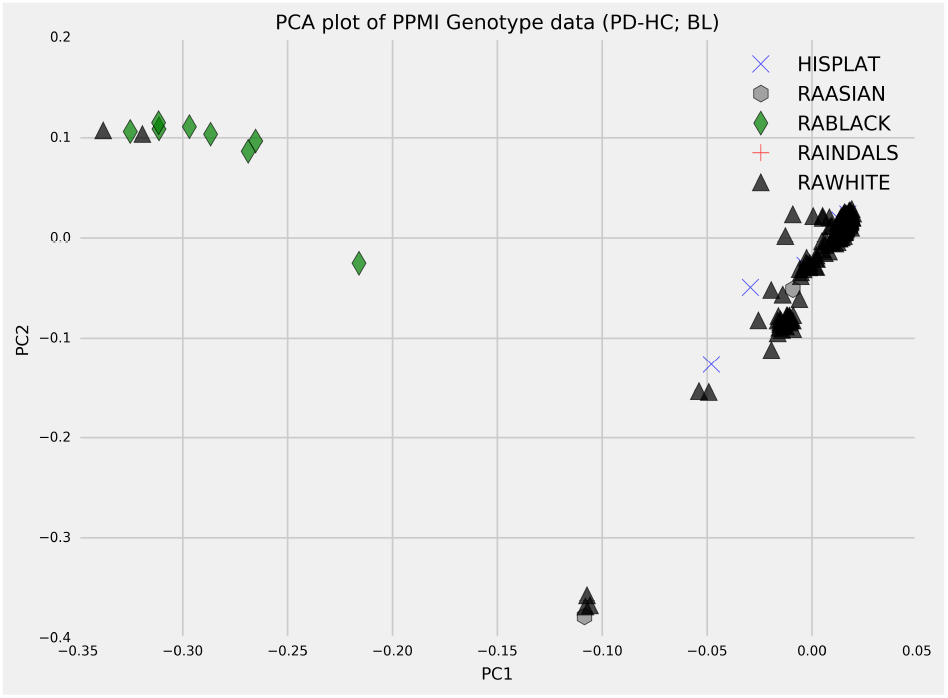
Scatterplot of the top two PCs computed on the genotype data reveals the population structure in PPMI data set (456 individuals).

**Figure 6:**
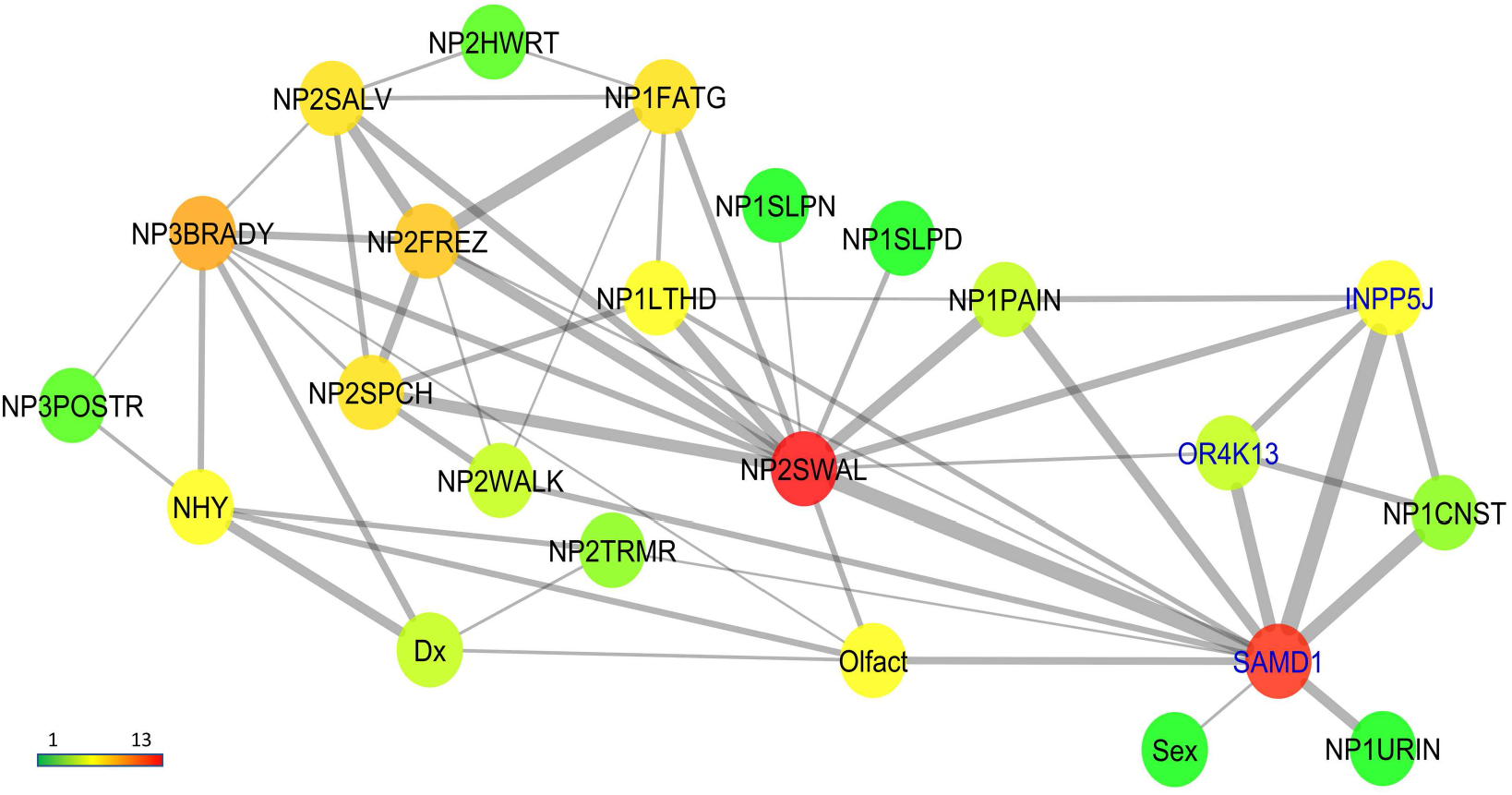
Interaction networks between genes and PD phenotypes with only top 20% of the edges present. Genes are highlighted in blue and phenotypes in black. The color of the nodes relate to their degrees, red being the highest and green being the lowest degree.

**Figure 7:**
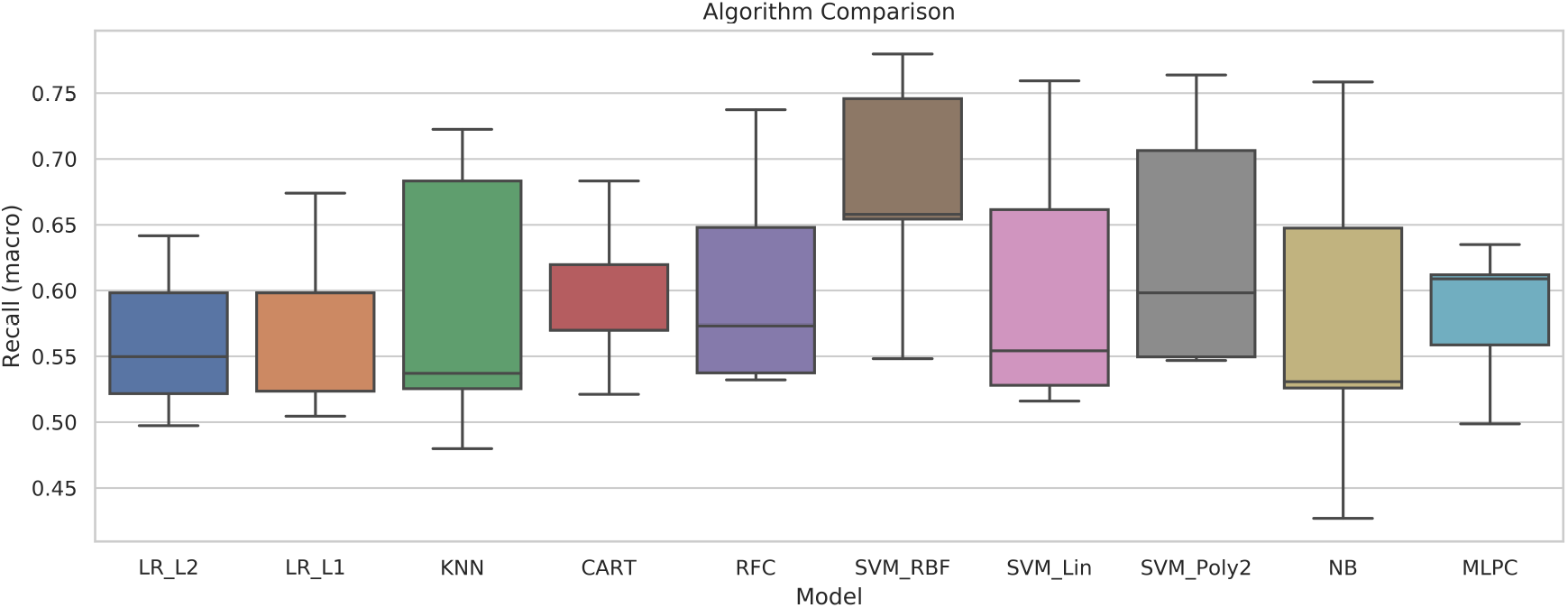
Classification performance comparison on training set with 24 eGenes and 75% of the 456 individuals (293 PD, 163 HC). We compare the Recall as we are more interested in the true positive classification and selected SVM with RBF kernel as it performed best.

**Table 2:** One *cis*-eSNP (other *cis*-eSNPs are not associated with expression in brain) and 18 protein-coding *cis-eGenes* highlighted by eQTL analysis (remaining 6 out of 24 *cis-eGenes* are pseudogenes). The tissues they are expressed in (GTEx v8), along with the reported regions in the brain are shown. Previously reported implications in PD are cited when available. ALL brain regions include: Amygdala (AM), Basal ganglia (BG), Cerebellum (CE), Cortex (CO), Cerebellar hemisphere (CH), Hippocampus (HI), Hypothalamus (HY), Spinal cord cervical (SCC), Substantia nigra (SN).

| Genetic variant | GTEx expression tissue | Brain Regions | Reported |
| --- | --- | --- | --- |
| rs60444836 | Brain | AM | - |
| NEDD4 | Brain | ALL | [29] |
| RNASE4 | Brain | ALL | - |
| SLC25A51 | Brain | ALL | - |
| LRTM1 | Brain | AM, Anterior cingulate cortex, BG, CO, Frontal cortex, HI | [32] |
| CEACAM8 | Brain | CH, CE, CO, SN | - |
| KIF18B | Brain | ALL | - |
| GPRC5B | Brain | ALL | [40] |
| AGO2 | Brain, Pancreas, Liver, Whole blood, Lung, Stomach, Kidney cortex, etc. | ALL | [30] |
| ROS1 | Brain | ALL | [41] |
| AC097721 | Brain | BG, HI, HY, SCC, SN | - |
| CTB-5506.12 | Brain | ALL | - |
| KIF1A | Brain | ALL | [31] |
| FAM225B | Brain, Prostate, Uterus, Ovary, Thyroid, etc. | ALL | - |
| SAMD1 | Brain, Whole blood, Liver, Kidney Cortex, Stomach, etc. | ALL | - |
| INPP5J | Brain, Adipose, Uterus, Whole blood, Ovary, Lung, Liver, Kidney cortex, Stomach, etc. | ALL | - |
| LRP6 | Brain, Whole blood, Skin, Ovary, Kidney cortex, etc. | ALL | [42] |
| ENY2 | Brain, Whole blood, Stomach, Liver, Lung, Kidney cortex, etc. | ALL | - |
| DPMI1 | Brain, Whole blood, Skin, Pancreas, Stomach, Thyroid, Liver, etc. | ALL | - |

**Figure 8:**
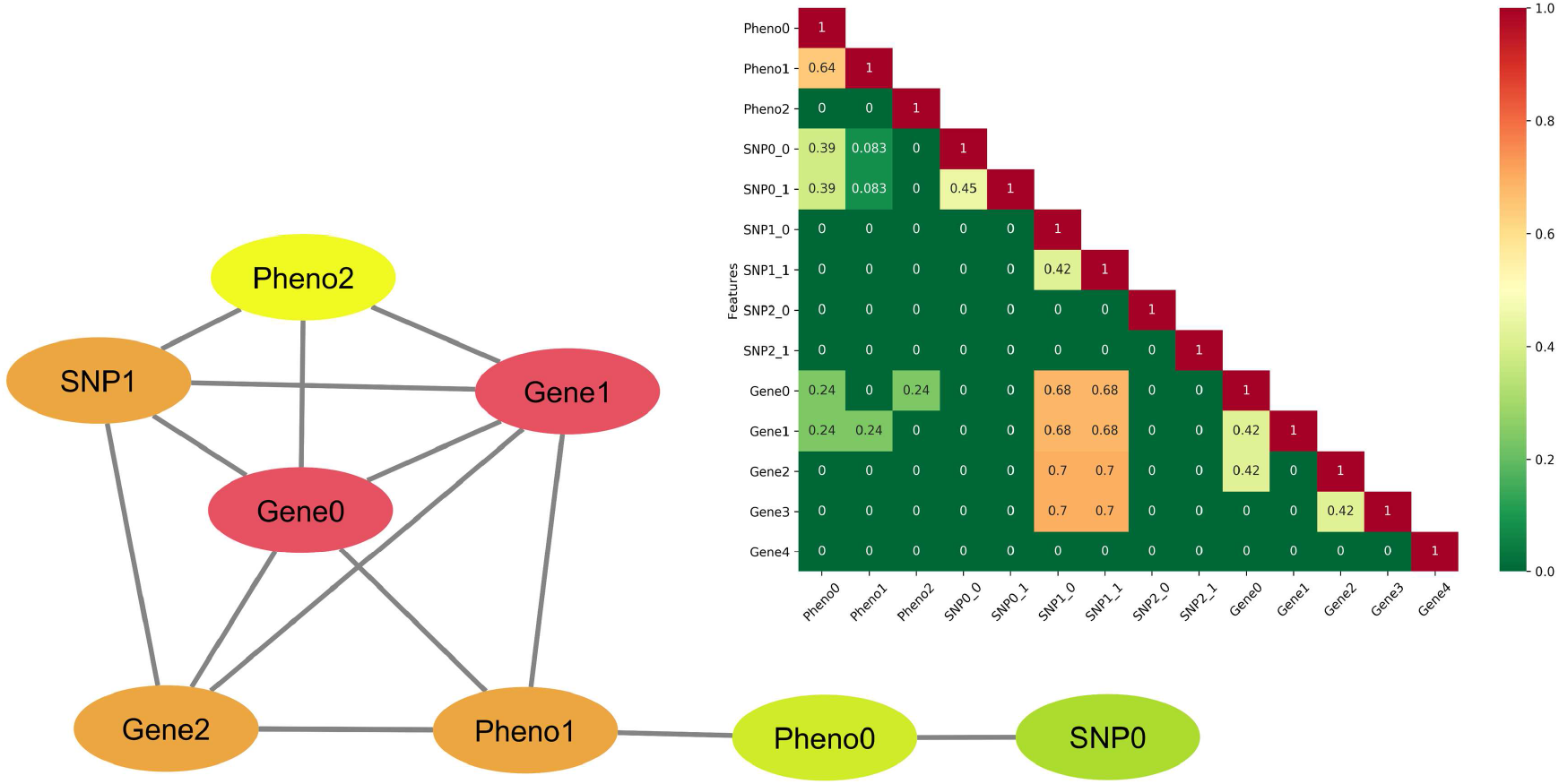
Network of the simulated variables colored by degrees of each node (darker colors have more degree). The correlation matrix of the variables is shown in the inset with the color gradient.

**Table 3:** *µ* for multivariate Gaussian distribution and standard variation *σ* for each parameter in the first simulation scenario.

| Features | $\mu$ | $\sigma$ |
| --- | --- | --- |
| Standard variation | 0.5 | 1.34 |
| Pheno1 | 0.5 | 1.16 |
| Pheno2 | -0.1 | 2 |
| SNP0 | 0.3 | 1.81 |
| SNP0 | 0.1 | 1.81 |
| SNP1 | 0.3 | 1.72 |
| SNP1 | 0.5 | 1.72 |
| SNP2.0 | 0.5 | 1 |
| SNP2.1 | 0.5 | 1 |
| Gene0 | 1 | 5.16 |
| Gene1 | 1 | 5.16 |
| Gene2 | 3 | 5.16 |
| Gene3 | 3 | 5.16 |
| Gene4 | 3 | 3 |

**Table 4:** Baseline proportions for binary variates computed from multivariate distributions corresponding to case-control proportions and MAFs in the first simulation scenario.

| $L$ | $P_L$ |
| --- | --- |
| Pheno0 | 0.667 |
| Pheno1 | 0.679 |
| Pheno2 | 0.48 |
| SNP0 | 0.588 |
| SNP1 | 0.53 |
| SNP2 | 0.59 |
| SNP1_1 | 0.648 |
| SNP2_0 | 0.691 |
| SNP2_1 | 0.691 |

**Table 5:**
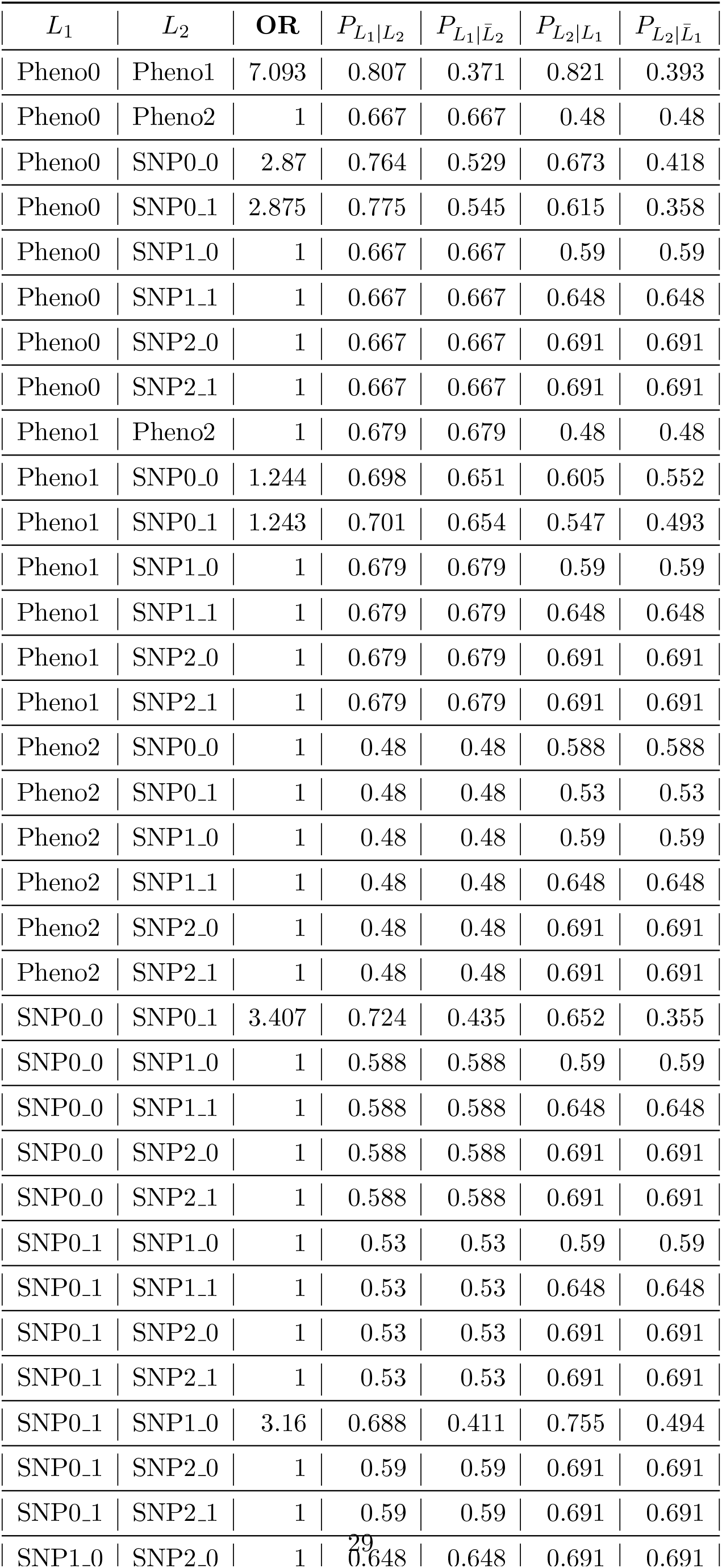
Odds ratios and proportions of binary measures given either phenotype state or allele in the first simulation scenario.

**Table 6:** *µ* for multivariate Gaussian distribution and standard variation *σ* for each parameter in the second simulation scenario.

| Features | mu | sigma |
| --- | --- | --- |
| Pheno0 | 0.5 | 2.28 |
| Pheno1 | 0.5 | 2.45 |
| Pheno2 | -0.1 | 2 |
| SNP0_0 | 0.3 | 1 |
| SNP0_1 | 0.1 | 1.81 |
| SNP1_0 | 0.3 | 1 |
| SNP1_1 | 0.5 | 1 |
| SNP2_0 | 0.5 | 1.64 |
| SNP2_1 | 0.5 | 1 |
| Gene0 | 1 | 5.43 |
| Gene1 | 1 | 5.43 |
| Gene2 | 3 | 3 |
| Gene3 | 3 | 7.86 |
| Gene4 | 3 | 3 |

**Table 7:** Baseline proportions for binary variates computed from multivariate distributions corresponding to case-control proportions and MAFs in the second simulation scenario.

| $L$ | $P_L$ |
| --- | --- |
| Pheno0 | 0.63 |
| Pheno1 | 0.625 |
| Pheno2 | 0.48 |
| SNP0_0 | 0.618 |
| SNP0_1 | 0.53 |
| SNP1_0 | 0.618 |
| SNP1_1 | 0.691 |
| SNP2_0 | 0.652 |
| SNP2_1 | 0.691 |

**Table 8:**
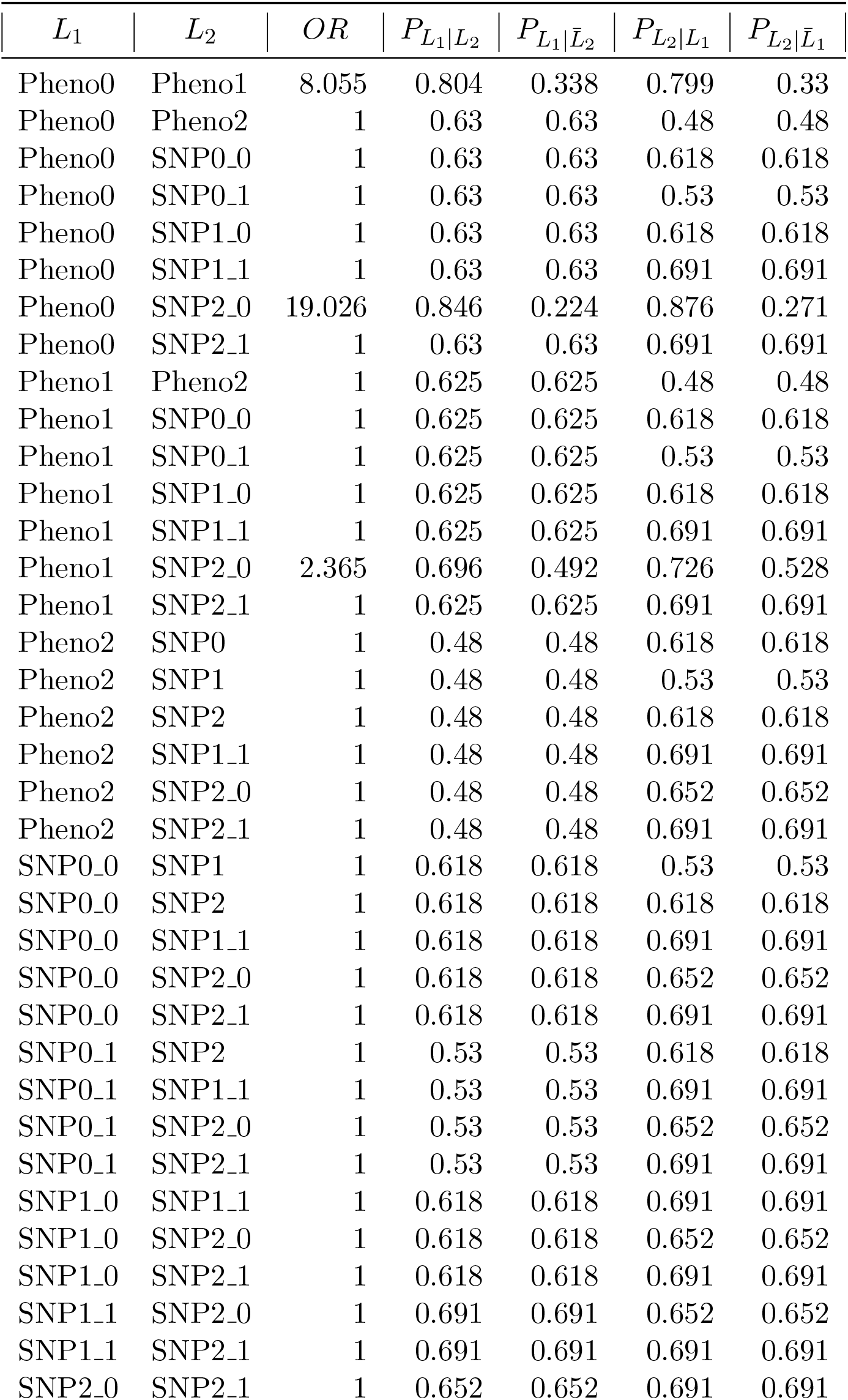
Odds ratios and proportions of binary measures given either phenotype state or allele in the second simulation scenario.

